# Proteomic associations with eating behaviors in young adults: a twin study

**DOI:** 10.64898/2026.04.14.26350850

**Authors:** Guiomar Masip, Gabin Drouard, Jaakko Kaprio

## Abstract

**Introduction:** Eating behaviors are consistently associated with weight-related traits, yet the biological factors contributing to individual differences in these behaviors remain poorly characterized. Plasma proteomics offers an opportunity to investigate the biological processes underlying eating behaviors.

**Methods:** Participants were 730 young adult twins from the FinnTwin12 cohort. Eating behaviors were measured through self-report questionnaires, including the Three-Factor Eating Questionnaire-R18 and four additional items on eating styles. Associations between plasma proteins and eating behaviors were examined using generalized estimating equation models adjusted for age and sex, with additional analyses adjusting for body mass index (BMI). Within-pair analyses were conducted in both monozygotic (MZ) and dizygotic twin pairs to assess whether associations were influenced by genetic or environmental factors.

**Results:** We identified 51 significant protein–eating behavior associations involving 35 unique proteins (FDR <0.05). We observed 19 associations for the item “overeating when feeling down” and 12 for the TFEQ factor of emotional eating. The identified proteins were predominantly enriched in immune system pathways, including the complement cascade and adaptive immune signaling. After further adjustment for BMI, 12 associations persisted, most of which were associated with eating-style items, suggesting that BMI had a substantial influence on protein–eating behavior associations. Within-pair analyses of MZ pairs indicated that several associations persist after accounting for genetic effects.

**Conclusion:** Our study identifies plasma proteins associated with eating behaviors, largely involving immune-related pathways. While some associations attenuated in twin analyses, several persisted, suggesting environmental influences. These results highlight potential biomarker candidates and indicate that modifiable environmental factors may contribute to the proteomic profiles associated with eating behaviors, with possible implications for weight-related traits.

## INTRODUCTION

Eating behaviors are key risk factors for eating-related disorders, including obesity and eating disorders (1). Overeating behaviors have been associated with obesity and binge-eating, whereas restrained eating behaviors have been associated with anorexia and bulimia nervosa (1).

The Behavioral Susceptibility Theory proposes that genetic susceptibility to obesity is expressed through individual differences in appetite, which in turn are manifested in eating behaviors (2). Several mediation studies have suggested that eating behaviors mediate the genetic susceptibility to obesity (3). On the other hand, obesity susceptibility genes are associated with eating behaviors and these associations are mediated by obesity outcomes (4,5). Hence, it is suggested that there might be a genetic overlap between obesity and eating behaviors, likely because most body mass index (BMI) single nucleotide polymorphisms (SNPs) are expressed in the central nervous system (6). Considering that several BMI-SNPs are expressed in the central nervous system, particularly in appetite-regulating regions (7), these findings highlight the need to elucidate the biological pathways underlying eating behaviors.

Proteomics, defined as the comprehensive characterization of the human proteome, offers a powerful approach to uncover molecular processes, providing opportunities to identify biomarkers and to understand biological mechanisms underlying complex traits (8) such as eating behaviors. While, to our knowledge, eating behaviors have not yet been studied through a proteomics lens, evidence from the literature on obesity and anorexia nervosa is informative. In obesity, proteomic analyses have shown that BMI-associated proteins are enriched in lipid metabolism and inflammatory pathways (9), and that BMI influences proteins involved in appetite regulation (10). In anorexia nervosa, proteomic studies have identified proteins across multiple biological systems, with the most consistent findings in metabolic and immune pathway (11). These studies illustrate the potential of proteomics to reveal systemic processes in weight-related disorders, highlighting its potential for advancing the study of eating behaviors. Consequently, identifying proteins associated with eating behaviors may pave the way for a better understanding of the biological underpinnings of these conditions.

Blood proteomic variation is heterogeneously shaped by both genetic and environmental factors (12), with implications for understanding causal roles and potential modifiability (13). Proteins associated with eating behaviors primarily due to environmental factors may be good candidates for intervention, as public health actions and health professionals can potentially modify them, whereas associations driven by genetic factors may be more useful for the early identification of at-risk individuals. However, while identifying genetic associations is possible in studies of unrelated individuals, fully ruling out genetic effects may be challenging. The use of twins and other family-based designs would enable a broader understanding of the role of genetics and the environment determinants in protein-eating behavior associations.

Our study aims were to identify plasma proteins associated with individual differences in eating behaviors, characterize the biological pathways underlying these behaviors, and determine whether identified associations persist independently of genetic factors. To do so, we leveraged data from 730 young adult twins for whom comprehensive proteomic and eating behavior data were available.

## METHODS

### Participants

The data for this study were derived from the population-based study FinnTwin12 (FT12), which aims to investigate behavioral development and health habits in adolescents and young adults (14). Within this cohort, an intensive subset of twins underwent semi-structured psychiatric interviews, additional questionnaires, and collection of venous blood samples when they turned adults (age range: 21-25), from which plasma proteomics were obtained. At the time of blood sampling, the twins’ weight and height were measured, from which BMI was calculated (kg/m^2^).

In this study, we excluded participants who were pregnant or on cholesterol medication. The final sample comprised 730 twin individuals, including 123 monozygotic (MZ) and 101 same-sex dizygotic (DZ) complete twin pairs. All had plasma proteomics data and had also completed the eating behaviors and dietary habits questionnaires. The data collection was approved by the Ethics Committee of the Helsinki and Uusimaa University Hospital District. Written informed consent was obtained from all participants.

### Assessment of eating behaviors

Eating behaviors were assessed using the 18-item Three-Factor Eating Questionnaire (TFEQ-R18), which assesses current dietary practices across three dimensions: restrained eating, uncontrolled eating and emotional eating. Each item is rated on a 4-point Likert scale (usually to seldom), and the scores were summed up to obtain total scores for each dimension, as described previously (15,16), i.e. cognitive restraint, uncontrolled eating, and emotional eating.

Furthermore, participants were asked to choose one of four options (usually to seldom) that best described their overall eating style: one item for low meal frequency, one item for regular eating, one item about eating at the dining facility and one item for alternating restriction and overeating style.

### Proteomic data processing

Proteomic data from the twins’ plasma samples were generated in four batches using LC-ESI-MS/MS (Q Exactive HF mass spectrometer) at the Turku Proteomics Facility (Finland), as described elsewhere (17). Plasma proteins were processed following the standard workflow of the Turku Proteomics Facility, including depletion of the 14 most abundant proteins using a commercial kit (High Select™ Top14 Abundant Protein Depletion Mini Spin Columns, Thermo Scientific, cat. No: A36370). Protein digestion, peptide desalting, and subsequent liquid chromatography–mass spectrometry (LC– MS/MS) analysis were also carried out. Spectronaut software was used for protein identification and quantification, with local data normalization applied. Data processing of raw protein values has been described in detail elsewhere (18). Briefly, raw values were log_2_-transformed, and proteins with more than 10% missing data were excluded. Remaining missing values were imputed using the lowest observed value per batch, and batch effects were corrected using Combat (19). The resulting dataset included 439 proteins, and proteins were scaled to a mean of zero and variance one.

### Statistical analysis

We assessed the associations between eating behavior variables and blood proteins using generalized estimating equation (GEE) models. These models account for the data structure, making them suitable for twin data by correcting for family relatedness. Proteins were fitted as dependent variables and eating behavior variables — either eating behavior items and factors and eating styles items — were fitted as independent variables. Sex and age at blood sampling were included as covariates, with BMI additionally adjusted for in subsequent analyses. We tested the null hypothesis of no association for eating behavior variables and corrected the resulting p-values for multiple testing using the Benjamini-Hochberg method in each set of analysis (i.e., for each eating behavior), considering associations significant at FDR < 0.05. GEE models were fitted using the *geepack* R package (version 1.3.9).

For the identified statistically significant associations in the individual-based analyses, we aimed to assess whether they persisted when correcting for genetic confounding. To this end, we examined MZ and DZ twin pairs. As MZ twins share their entire DNA sequence, differences in their traits (whether protein levels or eating behaviors) are expected to reflect environmental influences, in the broadest sense possible. Conversely, DZ twins share on average half of their segregating genes; differences within DZ pairs may therefore still originate from genetic effects. We therefore performed within-pair analyses in two steps. First, we ran within-pair analyses combining MZ and DZ pairs to account for shared environmental influences and only part of the genetic factors while maximizing statistical power. Then, we restricted the analyses to MZ twin pairs to fully control for genetic effects. We then performed linear regression models, fitting differences in protein levels as the dependent variable and differences in eating behaviors as the independent variable, restricted to the associations identified in the previous step.

## RESULTS

General characteristics of the participants, including age, sex, BMI, the TFEQ-R18 factors, and all eating behavior and eating-style items, are presented in **Table 1**.

**Table 1.** General characteristics of the study sample.

|  | N | All | N | Males | N | Females |
| --- | --- | --- | --- | --- | --- | --- |
| Age, years, mean (SD) | 729 | 22.33 (0.63) | 320 | 22.38 (0.69) | 409 | 22.29 (0.57) |
| Body mass index, kg/m<sup>2</sup>, mean (SD) | 729 | 23.31 (3.91) | 320 | 23.89 (3.73) | 409 | 22.86 (3.99) |
| ***TFEQ-R18 items*** |  |  |  |  |  |  |
| Difficulty resisting tempting foods, mean (SD) | 724 | 3.06 (0.87) | 319 | 2.89 (0.88) | 405 | 3.20 (0.84) |
| Portion size control, mean (SD) | 725 | 1.84 (0.80) | 319 | 1.52 (0.64) | 406 | 2.09 (0.83) |
| Eating when feeling nervous or agitated, mean (SD) | 723 | 3.13 (0.83) | 319 | 3.33 (0.74) | 404 | 2.97 (0.86) |
| Difficulty stopping eating, mean (SD) | 724 | 3.23 (0.86) | 319 | 3.40 (0.78) | 405 | 3.10 (0.89) |
| Social eating influence, mean (SD) | 725 | 2.66 (0.76) | 319 | 2.71 (0.78) | 406 | 2.63 (0.75) |
| Overeating when feeling down, mean (SD) | 721 | 3.07 (0.91) | 317 | 3.48 (0.69) | 404 | 2.75 (0.94) |
| Eating in response to food cues, mean (SD) | 722 | 2.88 (0.79) | 317 | 2.97 (0.80) | 405 | 2.81 (0.77) |
| Extreme hunger sensations, mean (SD) | 725 | 3.03 (0.81) | 319 | 3.03 (0.82) | 406 | 3.03 (0.79) |
| Difficulty stopping eating until the plate is finished, mean (SD) | 722 | 3.08 (0.80) | 317 | 2.97 (0.84) | 405 | 3.17 (0.75) |
| Eating to cope with loneliness, mean (SD) | 723 | 3.26 (0.85) | 318 | 3.65 (0.61) | 405 | 2.95 (0.89) |
| Meal intake control, mean (SD) | 725 | 1.97 (0.87) | 319 | 1.66 (0.75) | 406 | 2.21 (0.89) |
| Avoidance of energy-dense foods, mean (SD) | 724 | 2.01 (0.96) | 319 | 1.71 (0.79) | 405 | 2.25 (1.01) |
| Persistent feelings of hunger, mean (SD) | 724 | 2.75 (0.86) | 318 | 2.64 (0.90) | 406 | 2.83 (0.83) |
| Feeling hungry between meals, mean (SD) | 725 | 2.06 (0.67) | 319 | 2.10 (0.72) | 406 | 2.02 (0.62) |
| Avoidance of tempting foods, mean (SD) | 725 | 2.45 (0.81) | 319 | 2.22 (0.77) | 406 | 2.63 (0.79) |
| Eating less than desired, mean (SD) | 724 | 1.76 (0.82) | 319 | 1.55 (0.70) | 405 | 1.92 (0.86) |
| Eating large amounts when not hungry, mean (SD) | 723 | 2.03 (0.77) | 319 | 1.96 (0.75) | 404 | 2.09 (0.79) |
| Unrestricted eating, mean (SD) | 475 | 2.82 (0.93) | 240 | 2.68 (0.94) | 235 | 2.97 (0.89) |
| ***TFEQ-R18 factors*** |  |  |  |  |  |  |
| Emotional eating, mean (SD) | 725 | 28.34 (24.71) | 319 | 17.24 (19.01) | 406 | 37.07 (25.19) |
| Uncontrolled eating, mean (SD) | 725 | 34.78 (16.91) | 319 | 35.06 (17.55) | 406 | 34.57 (16.41) |
| Cognitive restraint, mean (SD) | 725 | 34.22 (20.80) | 319 | 25.68 (16.02) | 406 | 40.93 (21.67) |
| ***Eating-style items*** |  |  |  |  |  |  |
| Low meal frequency, mean (SD) | 717 | 2.19 (0.54) | 315 | 2.09 (0.54) | 402 | 2.27 (0.53) |
| Regular eating, mean (SD) | 718 | 2.41 (0.72) | 315 | 2.42 (0.75) | 403 | 2.41 (0.70) |
| Eating meals at the canteen, mean (SD) | 718 | 1.87 (1.11) | 315 | 1.74 (1.11) | 403 | 1.96 (1.10) |
| Alternating restriction and overeating style, mean (SD) | 717 | 1.47 (0.77) | 314 | 1.28 (0.57) | 403 | 1.63 (0.86) |

### Associations between protein abundance and eating behaviors

Associations between plasma proteins and eating behaviors, derived from models adjusted for sex and age, are presented in **Table 2**. We identified 51 significant associations at FDR <0.05, involving 35 unique proteins. Nine proteins were related to both the TFEQ-R18 subscale “overeating when feeling down” and the TFEQ-R18 factor emotional eating. Of the 51 associations, two were observed for the item of “portion size control”, one for “eating when feeling nervous”, 19 for “overeating when feeling down”, one for “extreme hunger sensation”, two for “meal intake control”, one for “eating less than desired” and one for “eating large amounts when not hungry”. For the TFEQ-R18 factors, we identified 15 associations: 12 with emotional eating and three with cognitive restraint. After additional adjustment for BMI, three associations remained significant: “eating when feeling nervous or agitated” and Heparanase (effect size = -0.16), “extreme hunger sensation” and Carboxypeptidase N subunit 2 (effect size = 0.15), and “eating large amounts when not feeling hungry” and Prostaglandin-H2 D-isomerase (effect size = -0.19).

**Table 2.**
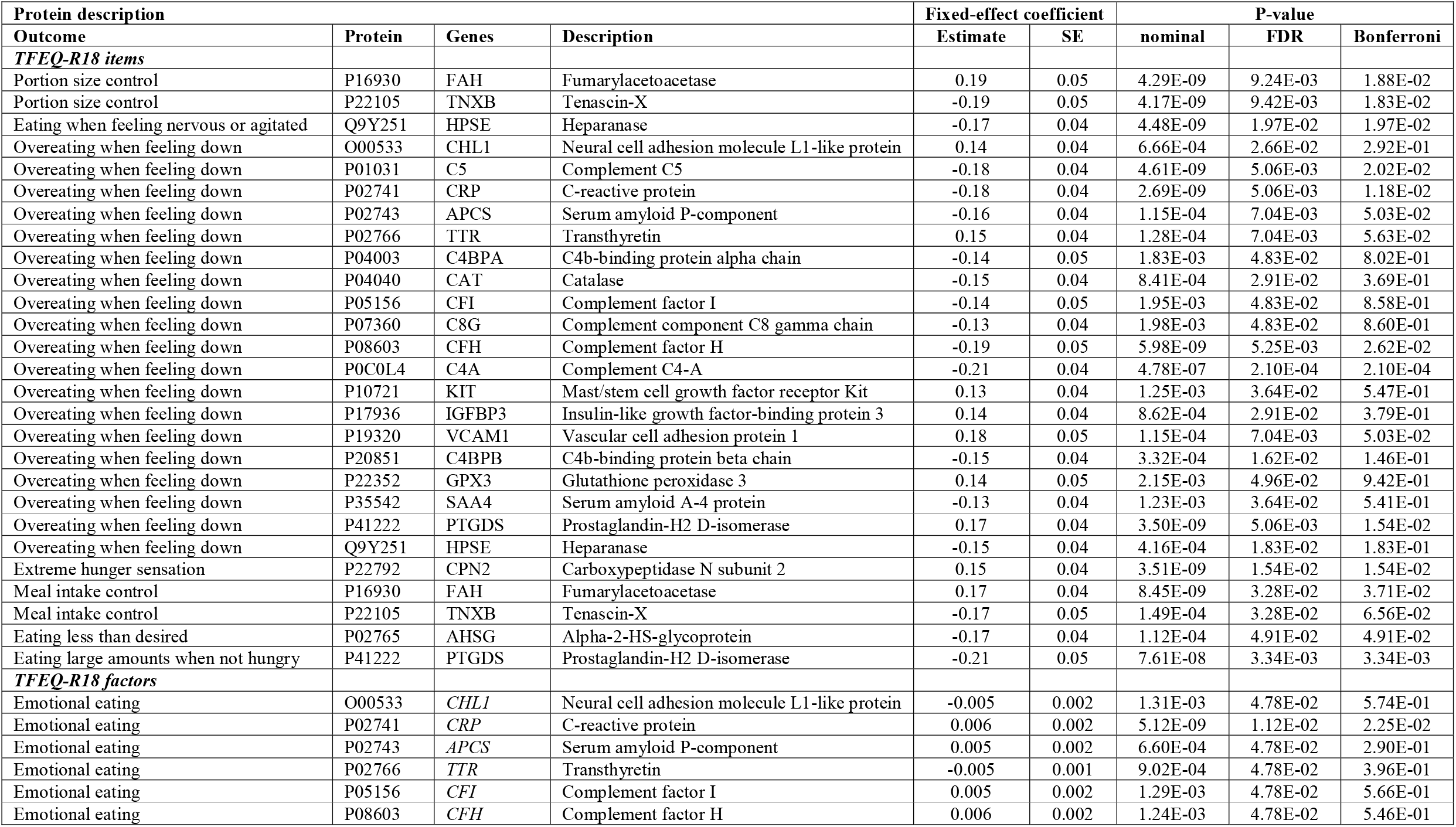
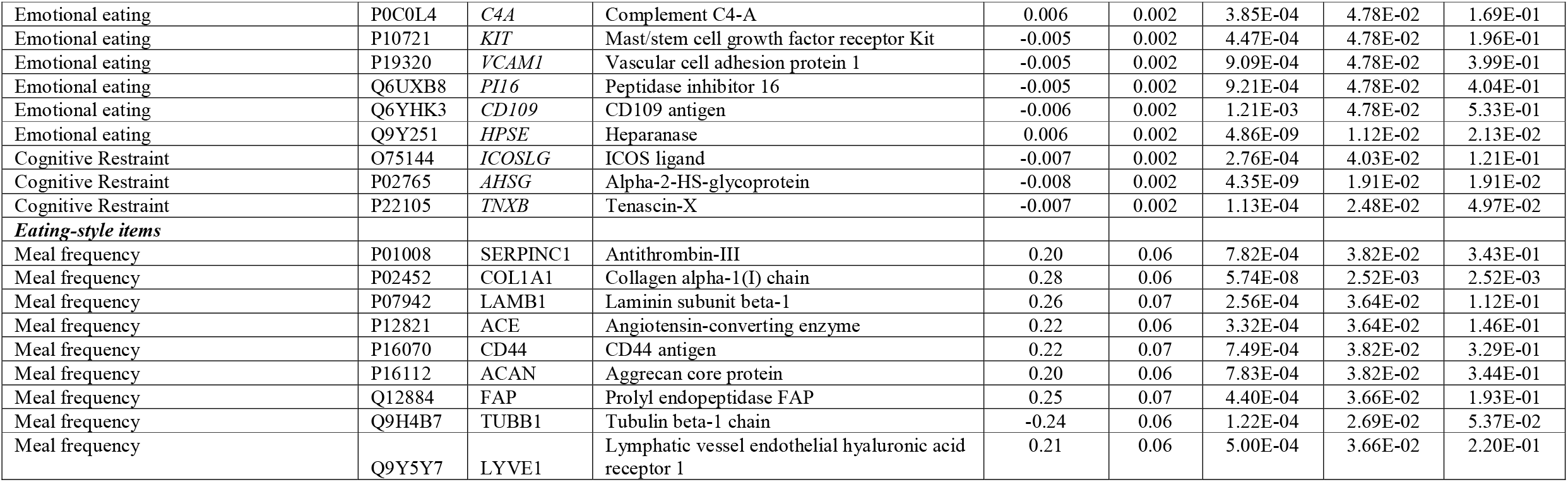
Associations between plasma protein abundance and eating behaviors.

For the eating-style items, associations adjusted for age and sex revealed nine significant associations for the “meal frequency” item. These associations persisted after further adjustment for BMI (**Table S1**). In addition, we observed two associations for the “regular eating” item and Angiogenin (effect size = 0.17) and Endoplasmic reticulum chaperone BiP (effect size = **-**0.18).

### Enriched biological processes and pathways

Pathway analyses of the 51 proteins associated with eating behaviors were conducted using the Reactome database. Significant pathways are presented in **Figure 1** and detailed pathway analyses are presented in **Table S2**. Enriched pathways were mainly related to the immune system, more specifically in the complement cascade and the adaptative immune signaling. Proteins were also enriched in other pathways including developmental biology, involving processes in nervous system development and pancreatic cell lineage differentiation, and extracellular matrix organization, encompassing collagen formation and integrin-mediated cell interactions. Additionally, several proteins were involved in protein metabolism, particularly in post-translational modification and amyloid fiber formation. Other pathways were also identified but were based on fewer entities, including those related to hemostasis, metabolism of carbohydrates or signal transduction.

**Figure 1.**
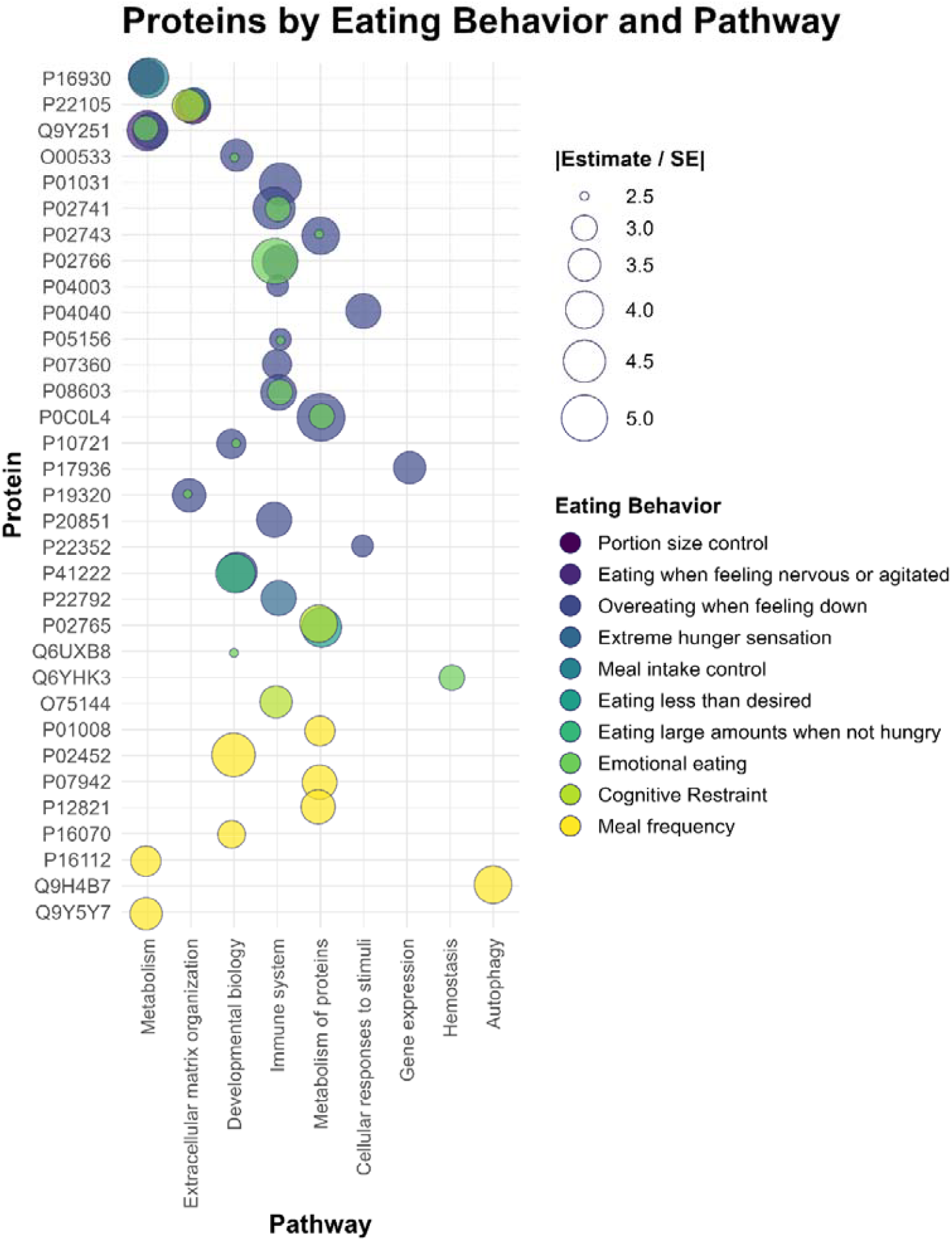
Plasma proteomics of eating behaviors and their enriched biological pathways. Bubble size represents the magnitude of the standardized effect estimate (Estimate / Standard Error). Two proteins with UniProt IDs P35542 and Q12884 were excluded due to the absence of pathway annotation in Reactome.

### Within-pair associations between protein abundance and eating behaviors

Analyses in all twin pairs pooled together (both MZ and DZ) revealed 30 of the 51 identified associations had FDR p-values <0.05 (**Table S3**). These associations therefore persist independently of environmental influences shared by co-twins. In analyses restricted to MZ pairs (**Table S3**), we observed that 21 associations had nominal p-values <0.05 and 18 associations had FDR p-values <0.05. This indicates that these associations persist independently of genetic factors, suggesting the influence of environmental factors and/or the possibility of a causal relationship.

## DISCUSSION

In this study involving young Finnish adults, we identified 51 associations between plasma proteins and eating behaviors, mainly involving pathways related to the immune system, protein metabolism and extracellular matrix organization. However, after adjusting for BMI, approximately three-fourths of identified associations did not pass multiple testing correction, suggesting a substantial role of BMI in protein-eating behavior associations. Furthermore, many associations persisted in within-pair analyses, which indicates genetic effects are not solely driving reported associations. To our knowledge, this study is the first to examine plasma proteins and eating behaviors, bridging a molecular gap between behavioral traits and biological processes, which may serve as a valuable resource for the scientific community.

The immune system was the most enriched pathway in this study, involving processes within the complement cascade. This finding is consistent with the fact that appetite is controlled in the central nervous system (20), and the complement system itself plays dual roles in both central nervous and immune systems (21). Previous studies have observed that inflammatory markers, such as higher C-reactive protein levels, are associated with changes in appetite and reduced food intake (22–24). Moreover, animal models support this association, showing that the complement factor C5a may promote food intake via the central nervous system (25).

This may reflect the interplay between central and peripheral mechanisms regulating appetite, as the complement system plays roles in both the central nervous and immune systems (21). Beyond immune-related pathways, we observed proteins enriched in metabolic and cellular structural processes. Although not directly comparable, similar biological pathways have been observed in proteomic studies of obesity (9) and anorexia nervosa (11), suggesting partially shared biological mechanisms. Among the 35 unique proteins identified in our study, 15 were also associated with BMI in a previous multi-omic analysis of the same FT12 cohort, with most of them enriched in the immune system (18). Given the relationship between emotional eating and depressive symptoms (26), we compared our results with another proteomic study of depressive symptoms in the FT12 cohort (27). However, we only identified five overlapping proteins: Tenascin-X, Serum amyloid P-component, Complement C4-A, Mast/stem cell growth factor receptor kit, and Insulin-like growth factor-binding protein-3, which were primarily involved in metabolic and cellular structural processes.

Hence, studies using the FT12 cohort suggest that eating behaviors may share a closer biological basis with obesity rather than with mood as indexed by depressive symptoms, although further studies are needed to assess generalizability to broader populations.

Our findings also highlight the relationship between eating behaviors and BMI. After adjusting for BMI, most of the associations between plasma proteins and eating behaviors measured with the TFEQ-R18 were attenuated. Obesity is associated with systemic inflammation and metabolic alterations that can affect central nervous system structure and function. Neuroimaging studies have shown that higher adiposity is associated with changes in gray and white matter in brain regions involved in appetite regulation and reward processes (28,29). In this study, we observed BMI to play a role in the association between plasma proteins and eating behaviors, with BMI potentially acting as a mediator or confounder, although longitudinal and mechanistic studies are needed to clarify causal pathways. The observed attenuation in coefficient magnitude reflects the complex interplay observed in previous genetic mediation studies of obesity and eating behaviors (1,3). Interestingly, associations between meal frequency and plasma proteins were robust to BMI adjustment, with all nine proteins remaining significant. Moreover, BMI-adjusted models revealed two additional associations for the eating-style item regular eating, involving Angiogenin and Endoplasmic reticulum chaperone BiP proteins. Previous studies have reported heterogeneous findings regarding eating styles and anthropometric measures, but it has been suggested that these associations might be more influenced by overall diet quality than by eating frequency or regularity (30–33). This may also explain the persistence of associations between eating styles and protein abundance after BMI adjustment, as eating styles reflect habitual and structural patterns of eating.

Several proteins remained significant in within-pair analyses. In MZ twin pairs only, we identified 21 significant associations between plasma proteins and the item overeating when feeling down and the emotional eating factor. This suggests that these associations do not arise solely from factors shared between co-twins, whether genetic or environmental, if such factors play any role at all. Significant findings in MZ twin pairs may also suggest a plausible causal relationship. Although three of these associations did not pass multiple testing corrections, the overall pattern of results points towards environmental exposures likely to modify key biological pathways and their influence on eating behaviors, which further studies are warranted to validate.

Despite an innovative study design, we acknowledge several potential limitations. Our study used a proteomic panel covering a limited subset of the entire proteome, which may have restricted our ability to identify all proteins relevant to eating behaviors. Further, proteomic platform differences may hinder replicability of our findings. Moreover, the relatively small sample size, particularly in within-pair analyses, together with the multiple testing correction, may have reduced our ability to detect further associations. That said, modest-to-large samples of twins or siblings with proteomic data are rare. Eating behaviors were obtained through self-reported questionnaires, and as with all studies relying on self-reported data, misreporting might have occurred, as individuals tend to provide the most socially desirable responses (34). Finally, our study is cross-sectional, thus we cannot draw any firm causal conclusions. However, within-pair modeling, particularly in MZ twin pairs, can be seen as a powerful approach to uncover plausible causal relationships, as a substantial proportion of confounding effects is controlled for by design.

Our study has important strengths. To our knowledge, this is one of the first studies, if not the first, to comprehensively investigate plasma protein associations with eating behaviors. We included eating styles in addition to eating behaviors to capture a broader picture of the whole eating process. This comprehensive approach helps to identify different biological and environmental factors that could contribute to various eating patterns and ultimately affect overall health. Furthermore, the inclusion of twin-pair analyses allowed for controlling genetic and shared environmental effects, providing insights into the relative contributions of these factors to eating behaviors.

In conclusion, this study identified several plasma proteins associated with eating behaviors in Finnish young adults, most of which were involved in immune-related pathways. These findings refine our understanding of the biological processes underlying individual differences in eating behaviors. Although some associations vanished in twin analyses, several associations persisted, suggesting that environmental factors contribute substantially to proteomic associations with eating behaviors, thus opening the door towards building tailored interventions. Future studies with longitudinal designs, ideally integrating multi-omics approaches, are needed to extend and validate these findings and to clarify the biological mechanisms underlying eating behaviors and their potential overlap with weight-related traits.

## Data Availability

The data used in the analysis is deposited in the Biobank of the Finnish Institute for Health and Welfare (https://thl.fi/en/web/thl-biobank/forresearchers). It is available to researchers after written application and following the relevant Finnish legislation.

## Author contributions

Conceptualization: GM and GD; Methodology: GM, GD and JK; Data curation: GD; Formal analysis: GD; Writing original draft: GM; Funding acquisition: JK; Draft review and editing: GM, GD and JK.

